# Declining prevalence of antibody positivity to SARS-CoV-2: a community study of 365,000 adults

**DOI:** 10.1101/2020.10.26.20219725

**Authors:** Helen Ward, Graham Cooke, Christina Atchison, Matthew Whitaker, Joshua Elliott, Maya Moshe, Jonathan C Brown, Barney Flower, Anna Daunt, Kylie Ainslie, Deborah Ashby, Christl Donnelly, Steven Riley, Ara Darzi, Wendy Barclay, Paul Elliott, for the REACT study team

## Abstract

**Background:** The prevalence and persistence of antibodies following a peak SARS-CoV-2 infection provides insights into its spread in the community, the likelihood of reinfection and potential for some level of population immunity.

**Methods:** Prevalence of antibody positivity in England, UK (REACT2) with three cross-sectional surveys between late June and September 2020. 365104 adults used a self-administered lateral flow immunoassay (LFIA) test for IgG. A laboratory comparison of LFIA results to neutralization activity in panel of sera was performed.

**Results:** There were 17,576 positive tests over the three rounds. Antibody prevalence, adjusted for test characteristics and weighted to the adult population of England, declined from 6.0% [5.8, 6.1], to 4.8% [4.7, 5.0] and 4.4% [4.3, 4.5], a fall of 26.5% [-29.0, −23.8] over the three months of the study. There was a decline between rounds 1 and 3 in all age groups, with the highest prevalence of a positive result and smallest overall decline in positivity in the youngest age group (18-24 years: −14.9% [-21.6, −8.1]), and lowest prevalence and largest decline in the oldest group (75+ years: −39.0% [-50.8, −27.2]); there was no change in antibody positivity between rounds 1 and 3 in healthcare workers (+3.45% [-5.7, +12.7]).

The decline from rounds 1 to 3 was largest in those who did not report a history of COVID-19, (−64.0% [-75.6, −52.3]), compared to −22.3% ([-27.0, −17.7]) in those with SARS-CoV-2 infection confirmed on PCR.

**Discussion:** These findings provide evidence of variable waning in antibody positivity over time such that, at the start of the second wave of infection in England, only 4.4% of adults had detectable IgG antibodies using an LFIA. Antibody positivity was greater in those who reported a positive PCR and lower in older people and those with asymptomatic infection. These data suggest the possibility of decreasing population immunity and increasing risk of reinfection as detectable antibodies decline in the population.

## Background

National prevalence surveys of SARS-CoV-2 antibodies provide critical insight into the extent that a population has been exposed to infection and may inform understanding of the future course of the epidemic.^1^ Studies in Iceland^2^ and Spain^3^ found quite different levels of population antibody positivity, with evidence of durable antibody response over 4 months from time of infection seen in Iceland. Meanwhile, cohort studies have suggested that antibody levels in individuals may fall substantially with time after infection, influenced by factors such as the severity of initial illness, age and co-morbidities.^4–9^

Changes in population antibody prevalence over time will be a complex interaction between the incidence of new infections and waning of antibody levels in those previously infected. Sequential antibody prevalence surveys can offer insight into the durability of antibody responses, key to understanding how developing immunity may prevent reinfection and limit further spread in the population.

In England, there was a large and widespread outbreak in March and April 2020 leading to high levels of hospitalisation and deaths.^10^ A national lockdown with the closure of schools, universities, hospitality, all but essential retail, and advice to work from home and avoid non-essential travel, was introduced in late March with a marked reduction in new infections until late August 2020.^11^

We have used a home-based testing approach to survey the extent of antibody positivity in the population indicative of SARS-CoV-2 infection. The lateral flow immunoassay (LFIA) employed allows a snapshot of antibody prevalence. Our first national survey in England, carried out among 105,000 individuals in late June 2020, found 6% of the adult population had detectable antibodies. Since the LFIA has a threshold for detection of a positive result, a decline in antibody level in individuals who have been infected may at some point result in negative tests, that is when the antibody levels fall below the threshold. Thus the proportion of positive tests in sequential random population samples can be used as an indicator of antibody waning.

The time-concentrated nature of the first wave of the UK epidemic provides an opportunity for measuring changes in antibody positivity in the population to estimate waning, and to quantify how this varies by sociodemographic and clinical characteristics. We report here prevalence of detectable antibody across three rounds of surveys (REACT-2 study^12-14^) involving representative cross-sections of the population of England.

## Methods

We analysed data from three rounds of a serial cross-sectional study of adults in England, UK that were carried out between June and September 2020 (Table 1). The protocol has been published;^12^ briefly, these were random, non-overlapping community samples from the adult population 18 years and older, using a self-administered LFIA test at home.^12–15^ Invitations were sent to named individuals randomly selected from the NHS patient list which includes anyone registered with a General Practitioner in England and covers almost the entire population. We aimed for a sample size of 100,000 in rounds 1 and 2 and 150,000 in round 3 to obtain prevalence estimates at lower tier local authority level. Sample size calculations are provided in the protocol,^12^ and the number of invitations sent out was based on an assumed response rate of 36 to 38% based on previous surveys. Registration was closed after 125,000 people signed up in rounds 1 and 2, and after 195,000 in round 3. Across all three rounds, 37.7% of those invited registered, and 29.9% provided a valid (IgG positive or negative) result (Supplementary appendix table S1). The response rate declined slightly over the three rounds. Those who registered were posted a self-administered point-of-care LFIA test (Fortress Diagnostics, Northern Ireland) with written and video instructions. The sensitivity of finger-prick blood (self-read) for IgG antibodies was 84.4% (70.5, 93.5) in RT-PCR confirmed cases in healthcare workers, and specificity 98.6% (97.1, 99.4) in pre-pandemic sera.^16^ Participants completed a short registration questionnaire (online/telephone) and a further survey upon completion of their self-test. Survey instruments are available on the study website (https://www.imperial.ac.uk/medicine/research-and-impact/groups/react-study/).

**Table 1:** Prevalence of antibody positivity to SARS-CoV-2 using LFIA test over three study rounds from June to September. Adjusted for test characteristics, weighted to the age, sex, region, ethnicity, index of multiple deprivation of England population (see Supplementary Appendix for detail on weighting)

|  | *Total*<br>*antibody*<br>*positive* | *Total tests (with*<br>*valid results)* | *Crude prevalence %*<br>*[95% CI]* | *Adjusted & weighted<sup>1</sup>*<br>*prevalence % [95% CI]* |
| --- | --- | --- | --- | --- |
| *Round 1 (20 Jun - 13 July)* | 5544 | 99908 | 5.55 [5.41-5.69] | 5.96 [5.78-6.14] |
| *Round 2 (31 Jul – 13 Aug)* | 4995 | 105829 | 4.72 [4.59-4.85] | 4.83 [4.67-5.00] |
| *Round 3 (15 - 28 Sept)* | 7037 | 159367 | 4.42 [4.32-4.52] | 4.38 [4.25-4.51] |

The prevalence from each round was calculated as the proportion of individuals reporting a valid test result who had a positive IgG result, adjusted for test performance,^17^ and weighted at national level for age, sex, region, ethnicity and deprivation to the adult population of England (Supplementary Appendix section 1.2). Change in prevalence was calculated between each round and from the first to the third round, and reported at national, regional and local geographic area, plus by key sociodemographic and clinical characteristics. Epidemic curves were constructed retrospectively from information from participants with a positive antibody test who had reported the date of onset for a confirmed or possible case of COVID-19.

To establish the sensitivity of the LFIA in relation to titres of neutralising antibodies we performed live virus neutralization tests on 49 sera from health care workers at 21 days or more since confirmed RT-PCR diagnosis of SARS CoV2 infection.^16^ Each of the sera was tested in the laboratory with the Fortress LFIA. In addition, the ability of the sera to neutralise wild type SARS-CoV-2 virus was assessed by neutralisation assay on Vero-E6 cells. Heat-inactivated sera were serially diluted in assay diluent consisting of DMEM (Gibco, Thermo Fisher Scientific) with 1% penicillin-streptomycin (Thermo Fisher Scientific), 0.3% BSA fraction V (Thermo Fisher Scientific). Two-fold serial dilutions starting at 1:10 were incubated with 100 TCID50/well of SARS-CoV-2/England/IC19/2020 diluted in assay diluent for 1 hr at room temperature and transferred to 96-well plates pre-seeded with Vero-E6 cells. Serum dilutions were performed in duplicate. Plates were incubated at 37°C, 5% CO_2_ for 4 days before staining the monolayers for surviving cells by adding an equal volume of 2X crystal violet stain to wells for 1 hr. Plates were washed, wells were scored for cytopathic effect and a neutralisation titre calculated as the reciprocal of the highest serum dilution at which full virus neutralisation occurred.

Data were analysed using the statistical package R version 4.0.0.^18^

We obtained research ethics approval from the South Central-Berkshire B Research Ethics Committee (IRAS ID: 283787), and Medicines and Healthcare products Regulatory Agency approval for use of the LFIA for research purposes only. A REACT Public Advisory Group provides input into the design and conduct of the research.

## Results

Results were available for 99,908, 105,829 and 159,367 people over the three rounds, which took place approximately 12, 18 and 24 weeks after the peak of the epidemic in England in early April. There were 17,576 positive tests in total. National antibody prevalence, adjusted for test characteristics and weighted to the adult population of England, declined from 6.0% [5.8, 6.1], to 4.8% [4.7, 5.00] and 4.4% [4.3, 4.5], a fall of 26.3% [-29.0, −23.8] over the three rounds. (Table 1, Figure 1) The fall was larger between rounds 1 and 2 (19.0% [-21.8, −16.1]) than between 2 and 3 (−9.1% [-12.0, −6.2]).

**Figure 1:**
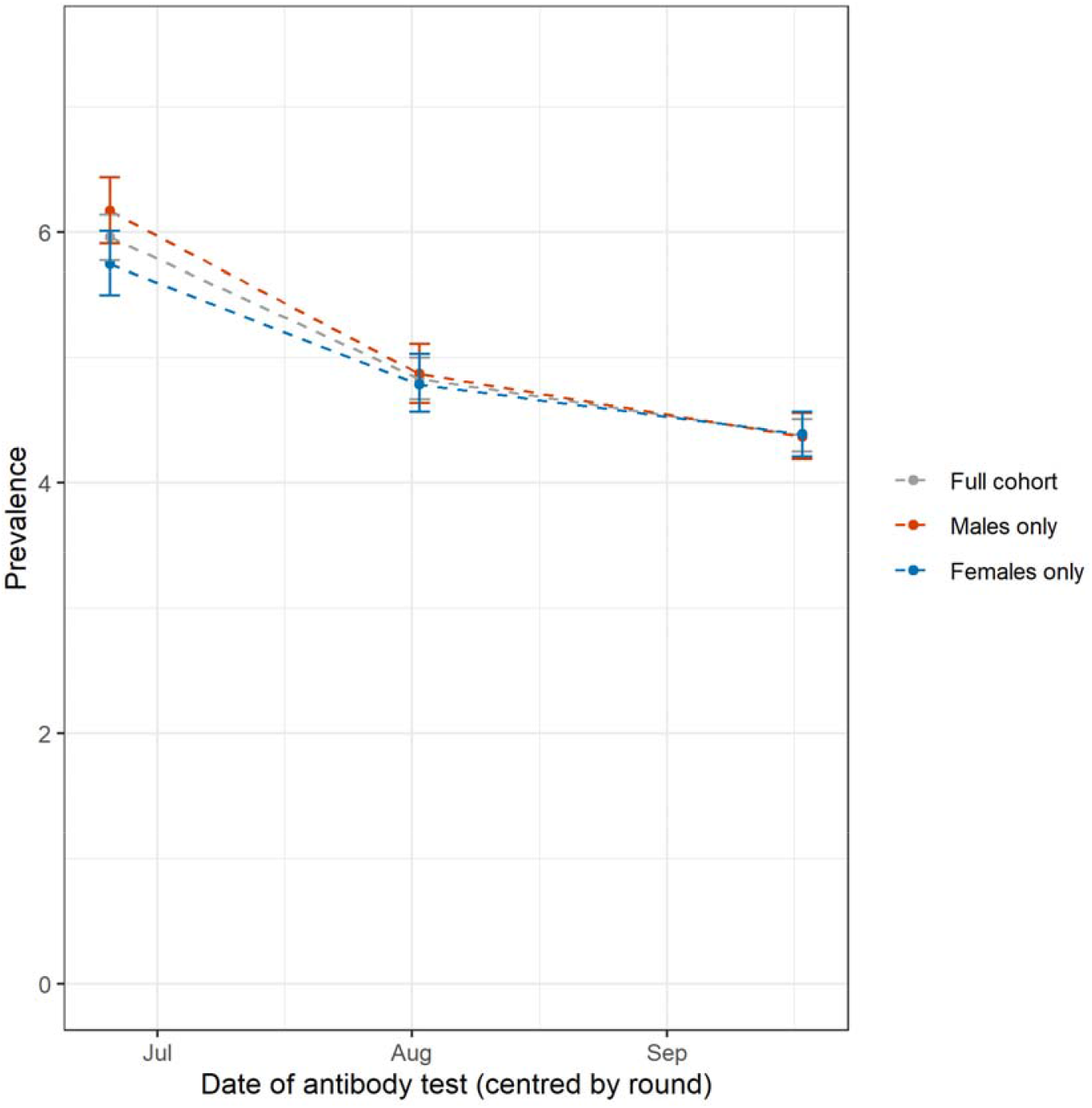
Prevalence of SARS-CoV-2 IgG antibody in England, by round of study (95% confidence intervals) for full cohort and by sex. Dates: Round 1 (June 20 – July 13 2020), Round 2 (31 July – 13 August 2020), Round 3 (15 - 28 September). Points show antibody prevalence by round of study. Prevalences are shown for the full sample (grey line), for male respondents only (red line) and for female respondents only (blue line). Error bars indicate 95% confidence intervals. Data points are aligned with the median response date within each round. All estimates of prevalence (95% confidence intervals) adjusted for imperfect test sensitivity and specificity, and re-weighted to account for sample design and for variation in response rate (age, sex, ethnicity, region and deprivation) to be representative of the England population (18+)

Over the three rounds of study we found similar patterns of infection to those reported in round 1^1^. Prevalence was highest for ages 18-24 years and lowest in those aged 75 and over. In the latest round, prevalence remained highest in London, at 9.5% (9.0, 9.9) compared with 1.6% (1.3, 1.9) in the South West of England; people of Black (includes Black Caribbean, African and Black British) and Asian (mainly South Asian) ethnicity had higher prevalence (13.8% [12.6-15.1] and 9.7% [9.1-10.4]) respectively, than those of white ethnicity (3.6% [3.5-3.8]). Prevalence was also higher among people working in health and social (residential) care, those living in more deprived areas and larger households (Table 2).

**Table 2:** Change in prevalence of antibody positivity to SARS-CoV-2 using LFIA test over three rounds from June to September. Adjusted and weighted (marked **, see Supplementary Appendix for methods) prevalence for each round by sociodemographic and clinical factors for each round of the study. Final columns show the percentage change (95% confidence limits) between R1 and R2, R2 and R3, and R1 and R3.

| Category | ROUND 1 |  |  | ROUND 2 |  |  | ROUND 3 |  |  | COMPARISONS |  |  |
| --- | --- | --- | --- | --- | --- | --- | --- | --- | --- | --- | --- | --- |
|  | Total antibody positive | Total tests (with valid results) | Prevalence R1 (weighted and adjusted where appropriate) | Total antibody positive | Total tests (with valid results) | Prevalence R2 (weighted and adjusted where appropriate) | Total antibody positive | Total tests (with valid results) | Prevalence R3 (weighted and adjusted where appropriate) | % difference R1/R2 | % difference R2/R3 | % difference R1/R3 |
| **Full cohort\*\*** |  |  |  |  |  |  |  |  |  |  |  |  |
| England | 5544 | 99908 | 5.96 [5.78-6.14] | 4995 | 105829 | 4.83 [4.67-5.00] | 7037 | 159367 | 4.38 [4.25-4.51] | -18.96 [-21.81, -16.11] | -9.11 [-12.01, -6.21] | -26.34 [-28.86, -23.83] |
| **Sex\*\*** |  |  |  |  |  |  |  |  |  |  |  |  |
| Male | 2405 | 43825 | 6.17 [5.91-6.44] | 2117 | 46269 | 4.87 [4.64-5.11] | 3029 | 69421 | 4.37 [4.19-4.56] | -21.07 [-24.96, -17.18] | -10.27 [-14.58, -5.95] | -29.17 [-32.74, -25.61] |
| Female | 3139 | 56083 | 5.75 [5.50-6.01] | 2878 | 59560 | 4.79 [4.57-5.03] | 4008 | 89944 | 4.39 [4.21-4.57] | -16.70 [-20.87, -12.52] | -8.56 [-12.73, -4.38] | -23.83 [-27.48, -20.17] |
| **Age\*\*** |  |  |  |  |  |  |  |  |  |  |  |  |
| 18-24 | 463 | 6499 | 7.86 [7.26-8.50] | 411 | 6493 | 7.31 [6.75-7.90] | 574 | 8763 | 6.70 [6.25-7.17] | -7.12 [-14.50, 0.25] | -8.34 [-15.32, -1.37] | -14.89 [-21.63, -8.14] |
| 25-34 | 930 | 13366 | 7.83 [7.35-8.32] | 775 | 13573 | 5.88 [5.47-6.32] | 1036 | 20212 | 5.15 [4.83-5.49] | -24.90 [-30.52, -19.28] | -12.24 [-18.54, -5.95] | -34.10 [-39.21, -28.99] |
| 35-44 | 964 | 17052 | 6.09 [5.65-6.56] | 837 | 17130 | 5.12 [4.71-5.55] | 1202 | 26687 | 4.60 [4.28-4.94] | -16.09 [-23.15, -9.03] | -10.16 [-17.38, -2.93] | -24.63 [-31.03, -18.23] |
| 45-54 | 1255 | 20634 | 6.41 [5.98-6.87] | 1100 | 21487 | 5.48 [5.08-5.90] | 1559 | 32403 | 4.96 [4.65-5.29] | -14.51 [-21.06, -7.96] | -9.49 [-16.06, -2.92] | -22.62 [-28.55, -16.69] |
| 55-64 | 1131 | 20404 | 5.92 [5.46-6.40] | 1074 | 21840 | 4.70 [4.30-5.14] | 1537 | 32870 | 4.33 [4.01-4.67] | -20.61 [-27.87, -13.34] | -7.66 [-15.53, 0.21] | -26.69 [-33.45, -19.93] |
| 65-74 | 568 | 15543 | 3.16 [2.76-3.59] | 594 | 17617 | 2.74 [2.37-3.14] | 787 | 26542 | 2.25 [1.96-2.56] | -13.61 [-25.95, -1.27] | -17.52 [-29.93, -5.11] | -28.80 [-39.87, -17.72] |
| 75+ | 233 | 6410 | 3.31 [2.86-3.79] | 204 | 7689 | 1.61 [1.26-2.00] | 342 | 11890 | 2.01 [1.70-2.34] | -51.06 [-63.44, -38.67] | 24.84 [3.73, 45.96] | -38.97 [-50.76, -27.19] |
| **Region\*\*** |  |  |  |  |  |  |  |  |  |  |  |  |
| North East | 196 | 3574 | 5.03 [4.30-5.85] | 202 | 4027 | 4.34 [3.66-5.10] | 296 | 6327 | 3.87 [3.33-4.46] | -13.92 [-28.43, 0.60] | -10.83 [-25.35, 3.69] | -23.26 [-36.38, -10.14] |
| North West | 714 | 11996 | 6.65 [6.14-7.19] | 657 | 12995 | 5.25 [4.80-5.74] | 910 | 18616 | 4.50 [4.15-4.87] | -21.05 [-28.42, -13.68] | -14.10 [-21.90, -6.29] | -32.18 [-38.80, -25.56] |
| Yorkshire and The Humber | 284 | 6519 | 3.95 [3.46-4.48] | 306 | 7391 | 3.97 [3.50-4.49] | 399 | 10594 | 3.37 [3.01-3.77] | 0.76 [-11.65, 13.16] | -15.37 [-26.20, -4.53] | -14.68 [-25.82, -3.54] |
| East Midlands | 601 | 12684 | 4.23 [3.71-4.80] | 533 | 13685 | 3.37 [2.90-3.89] | 756 | 20469 | 3.11 [2.73-3.52] | -20.33 [-32.39, -8.27] | -7.72 [-20.77, 5.34] | -26.48 [-37.35, -15.60] |
| West Midlands | 547 | 9620 | 5.82 [5.28-6.40] | 592 | 10062 | 6.97 [6.41-7.57] | 672 | 15046 | 4.77 [4.37-5.19] | 19.93 [10.31, 29.55] | -31.71 [-38.74, -24.68] | -18.04 [-26.29, -9.79] |
| East of England | 805 | 14433 | 5.09 [4.59-5.63] | 689 | 15189 | 4.02 [3.58-4.50] | 993 | 23174 | 3.69 [3.34-4.07] | -20.83 [-30.26, -11.39] | -8.21 [-18.41, 1.99] | -27.31 [-35.95, -18.66] |
| London | 1045 | 9547 | 12.96 [12.34-13.59] | 855 | 9872 | 9.38 [8.86-9.93] | 1265 | 15227 | 9.46 [9.03-9.91] | -27.55 [-31.94, -23.15] | 0.75 [-4.37, 5.86] | -27.01 [-31.10, -22.92] |
| South East | 995 | 21979 | 3.92 [3.54-4.32] | 891 | 22632 | 3.09 [2.75-3.45] | 1325 | 34738 | 3.01 [2.74-3.30] | -21.43 [-30.61, -12.24] | -2.27 [-12.30, 7.77] | -23.21 [-31.63, -14.80] |
| South West | 357 | 9556 | 2.79 [2.37-3.25] | 270 | 9976 | 1.28 [0.95-1.65] | 421 | 15176 | 1.62 [1.34-1.94] | -54.48 [-68.46, -40.50] | 27.34 [2.34, 52.34] | -41.94 [-55.20, -28.67] |
| **Employment** |  |  |  |  |  |  |  |  |  |  |  |  |
| Healthcare (patient-facing) | 379 | 3402 | 12.91 [11.61-14.32] | 389 | 3511 | 13.15 [11.87-14.52] | 578 | 5416 | 13.37 [12.33-14.47] | 1.70 [-8.37, 11.77] | 1.75 [-7.15, 10.65] | 3.49 [-5.73, 12.70] |
| Healthcare (other) | 73 | 1151 | 6.84 [5.15-8.91] | 62 | 1112 | 5.27 [3.74-7.20] | 113 | 1692 | 7.39 [5.95-9.07] | -22.95 [-48.54, 2.63] | 40.23 [9.87, 70.59] | 8.04 [-16.52, 32.60] |
| Care home (client-facing) | 115 | 761 | 19.56 [16.42-23.10] | 83 | 727 | 14.46 [11.67-17.72] | 108 | 979 | 11.09 [8.96-13.59] | -26.02 [-41.87, -10.17] | -23.24 [-41.36, -5.12] | -43.20 [-57.52, -28.89] |
| Care home (other) | 12 | 146 | 9.02 [4.53-16.26] | 12 | 224 | 3.63 [1.13-8.16] | 23 | 257 | 18.16 [13.28-24.23] | -59.76 [-110.98, -8.54] | 400.28 [277.96, 522.59] | 101.33 [40.47, 162.20] |
| Other essential worker | 1209 | 19927 | 6.63 [6.21-7.07] | 1019 | 19615 | 5.23 [4.85-5.64] | 1463 | 29572 | 4.93 [4.62-5.25] | -21.12 [-27.15, -15.08] | -5.74 [-12.43, 0.96] | -25.64 [-31.22, -20.06] |
| Other worker | 2189 | 37855 | 6.50 [6.20-6.82] | 1982 | 40782 | 5.17 [4.90-5.44] | 2704 | 60731 | 4.35 [4.14-4.56] | -20.62 [-24.92, -16.31] | -15.86 [-20.50, -11.22] | -33.23 [-37.23, -29.23] |
| Not in employment | 1516 | 35737 | 4.18 [3.92-4.45] | 1412 | 39030 | 3.38 [3.15-3.63] | 1988 | 59369 | 3.10 [2.91-3.29] | -18.90 [-24.64, -13.16] | -8.58 [-14.79, -2.37] | -25.84 [-31.10, -20.57] |
| **Resident in a care home** |  |  |  |  |  |  |  |  |  |  |  |  |
| Yes | 6 | 131 | 3.83 [0.86-9.92] | 18 | 259 | 6.69 [3.66-11.23] | 25 | 348 | 6.97 [4.23-10.83] | 74.67 [-27.94, 177.28] | 4.04 [-47.09, 55.16] | 81.72 [-15.67, 179.11] |
| No | 5538 | 99777 | 5.00 [4.83-5.17] | 4977 | 105570 | 3.99 [3.84-4.15] | 7012 | 159019 | 3.63 [3.51-3.75] | -20.20 [-23.40, -17.00] | -9.02 [-12.53, -5.51] | -27.40 [-30.20, -24.60] |
| **Ethnicity\*\*<sup>1</sup>** |  |  |  |  |  |  |  |  |  |  |  |  |
| White | 4827 | 92737 | 5.01 [4.83-5.19] | 4384 | 98003 | 4.05 [3.89-4.22] | 6176 | 148227 | 3.63 [3.50-3.76] | -19.16 [-22.55, -15.77] | -10.37 [-14.07, -6.67] | -27.54 [-30.74, -24.35] |
| Mixed | 106 | 1347 | 8.92 [7.09-11.08] | 76 | 1308 | 6.19 [4.66-8.05] | 108 | 1865 | 5.69 [4.46-7.16] | -30.61 [-50.78, -10.43] | -7.92 [-31.99, 16.16] | -36.10 [-54.71, -17.49] |
| Asian | 369 | 3658 | 11.86 [10.99-12.77] | 340 | 3930 | 11.23 [10.40-12.10] | 439 | 5518 | 9.70 [9.06-10.38] | -5.31 [-12.48, 1.85] | -13.62 [-20.21, -7.03] | -18.21 [-24.70, -11.72] |
| Black | 135 | 900 | 17.34 [15.75-19.05] | 102 | 936 | 11.92 [10.56-13.42] | 154 | 1304 | 13.77 [12.56-15.06] | -31.26 [-39.91, -22.61] | 15.52 [4.53, 26.51] | -20.59 [-28.78, -12.40] |
| Other | 79 | 762 | 12.28 [10.21-14.66] | 70 | 939 | 8.25 [6.50-10.33] | 98 | 1393 | 8.25 [6.77-9.95] | -32.82 [-49.27, -16.37] | -0.12 [-20.85, 20.61] | -32.90 [-48.21, -17.59] |
| **IMD quintile\*\*<sup>2</sup>** |  |  |  |  |  |  |  |  |  |  |  |  |
| Most deprived: 1 | 682 | 10082 | 7.28 [6.84-7.74] | 639 | 10997 | 6.27 [5.86-6.69] | 827 | 15681 | 5.39 [5.08-5.71] | -13.87 [-19.64, -8.10] | -14.04 [-19.78, -8.29] | -25.96 [-31.18, -20.74] |
| 2 | 947 | 16015 | 6.43 [6.03-6.85] | 855 | 16973 | 5.35 [4.98-5.73] | 1183 | 25206 | 4.82 [4.53-5.12] | -16.80 [-22.71, -10.89] | -9.91 [-16.07, -3.74] | -25.04 [-30.48, -19.60] |
| 3 | 1196 | 21474 | 5.87 [5.48-6.29] | 1027 | 23231 | 4.46 [4.12-4.82] | 1518 | 34548 | 4.37 [4.09-4.66] | -24.19 [-30.49, -17.89] | -2.02 [-8.97, 4.93] | -25.72 [-31.52, -19.93] |
| 4 | 1287 | 24840 | 5.22 [4.84-5.62] | 1182 | 25979 | 4.25 [3.91-4.61] | 1678 | 39595 | 3.84 [3.57-4.12] | -18.58 [-25.48, -11.69] | -9.65 [-16.94, -2.35] | -26.44 [-32.76, -20.11] |
| Least deprived: 5 | 1432 | 27497 | 4.99 [4.61-5.39] | 1292 | 28649 | 3.85 [3.52-4.21] | 1831 | 44337 | 3.51 [3.24-3.78] | -22.65 [-29.86, -15.43] | -9.09 [-16.88, -1.30] | -29.66 [-36.27, -23.05] |
| Household size |  |  |  |  |  |  |  |  |  |  |  |  |
| 1 | 720 | 15052 | 4.08 [3.68-4.50] | 671 | 16777 | 3.13 [2.79-3.50] | 953 | 24735 | 2.96 [2.67-3.25] | -23.04 [-32.11, -13.97] | -5.75 [-15.97, 4.47] | -27.45 [-36.03, -18.87] |
| 2 | 1784 | 36413 | 4.22 [3.95-4.49] | 1593 | 39252 | 3.20 [2.97-3.44] | 2273 | 59922 | 2.88 [2.70-3.07] | -24.17 [-30.09, -18.25] | -10.00 [-16.56, -3.44] | -31.75 [-36.97, -26.54] |
| 3 | 1158 | 19734 | 5.38 [5.00-5.79] | 1065 | 20898 | 4.45 [4.10-4.82] | 1484 | 31429 | 4.00 [3.73-4.29] | -17.29 [-24.16, -10.41] | -10.34 [-17.30, -3.37] | -25.84 [-31.97, -19.70] |
| 4 | 1204 | 19611 | 5.71 [5.32-6.13] | 1077 | 20110 | 4.77 [4.40-5.15] | 1549 | 30208 | 4.49 [4.20-4.80] | -16.46 [-23.12, -9.81] | -5.87 [-12.79, 1.05] | -21.37 [-27.50, -15.24] |
| 5 | 447 | 6403 | 6.72 [6.00-7.51] | 392 | 6174 | 5.96 [5.26-6.73] | 529 | 9354 | 5.13 [4.58-5.71] | -11.31 [-22.02, -0.60] | -13.93 [-24.66, -3.19] | -23.66 [-33.33, -13.99] |
| 6 | 152 | 1848 | 8.22 [6.82-9.84] | 124 | 1822 | 6.51 [5.23-8.02] | 162 | 2575 | 5.89 [4.84-7.10] | -20.80 [-38.08, -3.53] | -9.68 [-28.73, 9.37] | -28.47 [-44.28, -12.65] |
| 7+ | 79 | 827 | 9.82 [7.63-12.47] | 73 | 796 | 9.36 [7.18-12.02] | 87 | 1130 | 7.59 [5.88-9.64] | -4.68 [-28.62, 19.25] | -18.91 [-41.45, 3.63] | -22.71 [-44.09, -1.32] |
| Population density quintile |  |  |  |  |  |  |  |  |  |  |  |  |
| 1 | 808 | 19779 | 3.24 [2.91-3.58] | 680 | 20253 | 2.36 [2.07-2.67] | 964 | 30655 | 2.10 [1.87-2.34] | -27.16 [-36.73, -17.59] | -11.02 [-22.03, 0.00] | -35.19 [-43.83, -26.54] |
| 2 | 1002 | 19514 | 4.50 [4.14-4.88] | 894 | 20060 | 3.68 [3.35-4.04] | 1275 | 30773 | 3.31 [3.04-3.58] | -17.78 [-25.56, -10.00] | -10.60 [-18.75, -2.45] | -26.44 [-33.56, -19.33] |
| 3 | 1026 | 19817 | 4.55 [4.19-4.93] | 929 | 20090 | 3.88 [3.55-4.25] | 1258 | 30350 | 3.31 [3.04-3.58] | -14.73 [-22.42, -7.03] | -14.95 [-22.68, -7.22] | -27.47 [-34.51, -20.44] |
| 4 | 1145 | 20094 | 5.18 [4.80-5.58] | 950 | 20101 | 4.01 [3.66-4.37] | 1404 | 29967 | 3.96 [3.68-4.25] | -22.59 [-29.54, -15.64] | -1.25 [-8.98, 6.48] | -23.55 [-29.92, -17.18] |
| 5 | 1563 | 20704 | 7.41 [6.98-7.85] | 1542 | 25325 | 5.65 [5.30-6.01] | 2136 | 37622 | 5.15 [4.88-5.44] | -23.75 [-29.01, -18.49] | -8.67 [-14.16, -3.19] | -30.36 [-35.22, -25.51] |
| COVID history<sup>3</sup> |  |  |  |  |  |  |  |  |  |  |  |  |
| Positive test | 277 | 341 | 96.18 [90.78-100.00] | 274 | 361 | 89.76 [84.13-94.73] | 478 | 754 | 74.69 [70.48-78.74] | -6.67 [-11.89, -1.46] | -16.78 [-21.95, -11.61] | -22.33 [-26.98, -17.69] |
| Suspected by doctor | 353 | 1144 | 35.49 [32.35-38.79] | 348 | 1214 | 32.85 [29.87-35.99] | 470 | 1811 | 29.58 [27.21-32.07] | -7.41 [-16.03, 1.21] | -9.98 [-18.20, -1.77] | -16.65 [-24.49, -8.82] |
| Suspected by respondent | 3118 | 17893 | 19.31 [18.65-19.99] | 2742 | 16914 | 17.85 [17.19-18.52] | 3867 | 27546 | 15.23 [14.74-15.73] | -7.61 [-10.98, -4.25] | -14.62 [-17.82, -11.43] | -21.13 [-24.08, -18.18] |
| No | 1698 | 80390 | 0.86 [0.74-0.98] | 1565 | 87217 | 0.48 [0.37-0.58] | 2144 | 129126 | 0.31 [0.23-0.40] | -45.35 [-58.14, -32.56] | -33.33 [-52.08, -14.58] | -63.95 [-75.58, -52.33] |
| Symptom category<sup>4</sup> |  |  |  |  |  |  |  |  |  |  |  |  |
| No symptoms | 1791 | 81150 | 0.97 [0.85-1.10] | 1636 | 87971 | 0.55 [0.45-0.66] | 2267 | 130438 | 0.41 [0.32-0.49] | -44.33 [-55.67, -32.99] | -25.45 [-41.82, -9.09] | -58.76 [-69.07, -48.45] |
| Atypical symptoms only | 347 | 3426 | 10.52 [9.35-11.79] | 294 | 3185 | 9.43 [8.28-10.71] | 469 | 5108 | 9.38 [8.46-10.37] | -10.36 [-21.58, 0.86] | -0.64 [-11.88, 10.60] | -10.93 [-21.10, -0.76] |
| Screening symptoms | 3406 | 15332 | 25.08 [24.29-25.88] | 3065 | 14673 | 23.48 [22.70-24.28] | 4301 | 23821 | 20.07 [19.48-20.66] | -6.4 [-9.4, -3.3] | -14.5 [-17.4, -11.6] | -20.0 [-22.7, -17.3] |
| COVID contacts<sup>5</sup> |  |  |  |  |  |  |  |  |  |  |  |  |
| Yes, with confirmed case | 742 | 3946 | 20.97 [19.54-22.47] | 753 | 4543 | 18.28 [17.01-19.62] | 1107 | 7793 | 15.43 [14.52-16.38] | -12.78 [-19.22, -6.34] | -15.59 [-21.61, -9.57] | -26.37 [-32.09, -20.65] |
| Yes, with suspected case | 896 | 5307 | 18.65 [17.47-19.90] | 842 | 5115 | 18.15 [16.95-19.40] | 1121 | 7362 | 16.66 [15.69-17.67] | -2.73 [-9.12, 3.65] | -8.15 [-14.10, -2.20] | -10.67 [-16.46, -4.88] |
| No | 3906 | 90655 | 3.50 [3.35-3.67] | 3400 | 96171 | 2.57 [2.43-2.72] | 4809 | 144211 | 2.33 [2.22-2.44] | -26.57 [-30.86, -22.29] | -9.73 [-14.40, -5.06] | -33.71 [-37.43, -30.00] |

<sup>1</sup> Ethnicity categories: Asian includes Asian (south, east) and Asian British; Black includes Black African, Caribbean, Black British;
<sup>2</sup> Based on Index of Multiple Deprivation (2019) at lower super output area;
<sup>3</sup> COVID History is self-reported, based on response to the question, “Before you took this antibody test, did you think you had had COVID-19?” with response options of Yes, confirmed by a positive test (swab/\*PCR/antigen test); Yes, suspected by a doctor but not tested; Yes, my own suspicions; No.
<sup>4</sup> Symptom category is constructed from responses about self-reported specific symptoms. These were grouped into those reporting one or more “screening symptoms” based on recommendations for having a SARS-CoV-2 test (new persistent cough, fever, loss of sense of smell or taste), or atypical (any other symptom(s)), or none.
<sup>5</sup> Self reported contact with a case of COVID19

Table 2 and Figure 2 show the change in prevalence by round and overall by key covariates. There was a decline in prevalence between rounds 1 and 3 in all age groups, with the smallest overall decline at ages 18-24 years (−14.9% [-21.6, −8.1]) and largest at ages 75 years and over (−39.0% [-50.8, −27.2]). The decline from rounds 1 to 3 was largest in those who did not report a history of COVID-19, (−64.0% [-75.6, −52.3]), compared to −22.3% ([-27.0, −17.7]) in those with COVID-19 confirmed on PCR. There was no change in prevalence between rounds 1 and 3 in healthcare workers (+3.45% [-5.7, +12.7]).

**Figure 2.**
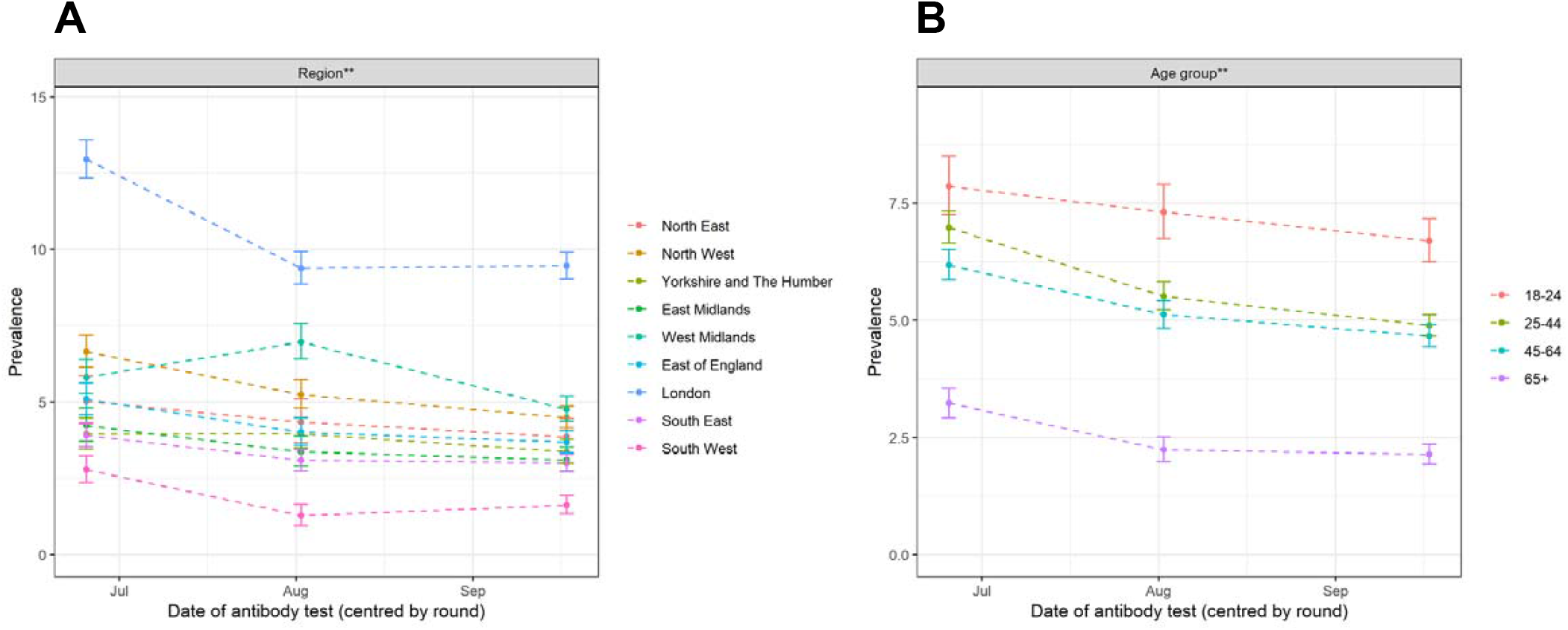
Antibody prevalence for each round of study, by (A) region and (B) age group, June to September 2020. A) Prevalence in each of the nine regions of England; B) prevalence by age group; Error bars indicate 95% confidence intervals. Prevalences are adjusted for known test performance and re-weighted where appropriate to be representative of the 18+ population of England (** denotes weighted prevalence). Survey responses were received across 2–3 week periods in each round (in late June, early August and mid-September); data points are aligned with the median response date within each round

Figure 3 shows how antibody prevalence changed between rounds at lower tier local area level (see also maps in Supplementary Appendix Figure 1). The slope of the fitted line approximates to the average decrease in prevalence, and the scatter shows the variation, with some areas seeing an increase and others a large decrease between rounds.

**Figure 3.**
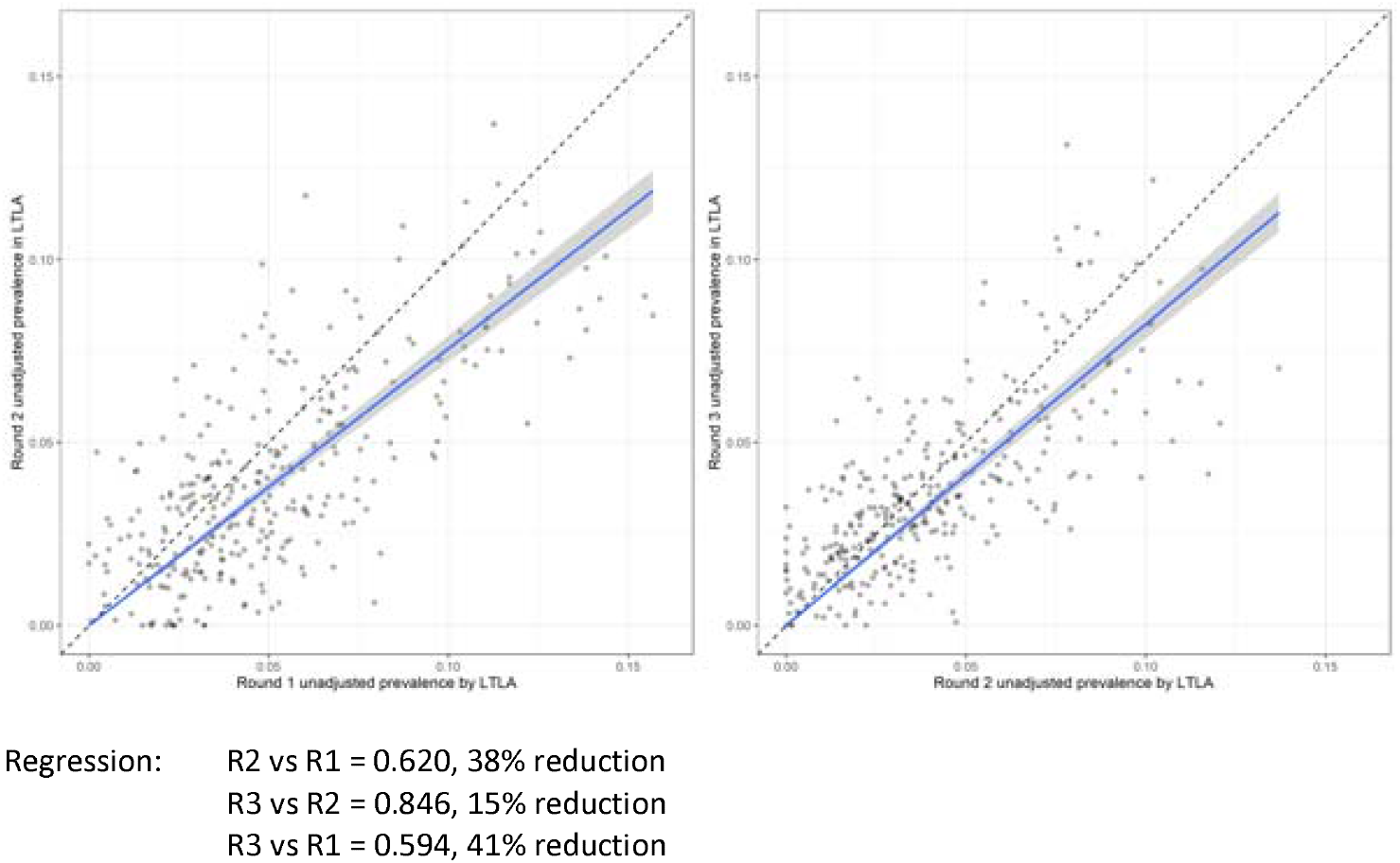
Antibody prevalence between rounds 1 and 2, and 2 and 3 by lower tier local authority. Scatterplot for each lower tier local authority showing change in prevalence from round 1 to round 2 (left) and round 2 to round 3 (right). The dashed line represents no change, the blue line the linear regression. The slope of the fitted line indicates the average decrease in prevalence, and the scatter shows the variation, with some areas seeing an increase and others a very large decrease between rounds.

The epidemic curves constructed from people who tested positive and reporting symptoms for each of the three rounds closely overlap, illustrating the relatively short, concentrated outbreak across the country with the majority of cases in March and April. (Figure 4) The figure also shows a steep decline in new cases from 6 April, 2 weeks after the national lockdown was introduced on 23 March. There was limited evidence of new cases after early May overall, but some apparent ongoing transmission in health and social care workers into May and June. (Figure 5). We noted a small increase in cases from late August and early September at the start of the second wave.

**Figure 4.**
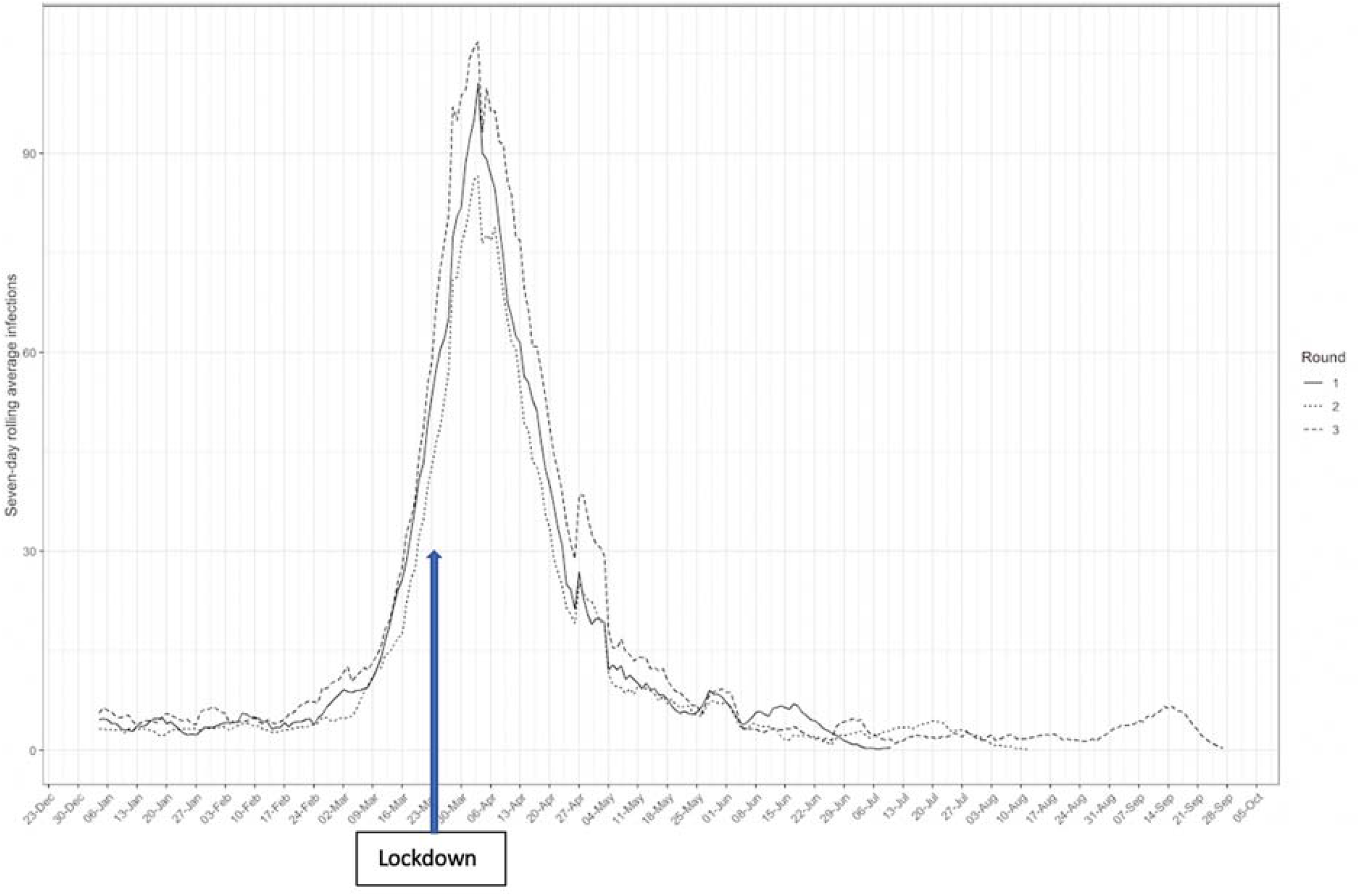
Epidemic curve reconstructed from reported date of onset from 17,576 IgG antibody positive people, by round of study. Seven-day rolling average of number of infections (by onset date) in 17,576 participants testing positive for antibodies and who reported a date of onset for symptoms of COVID19, shown separately for each round, together with an arrow indicating the date of the national lockdown in England (March 23^rd^ 2020).

**Figure 5.**
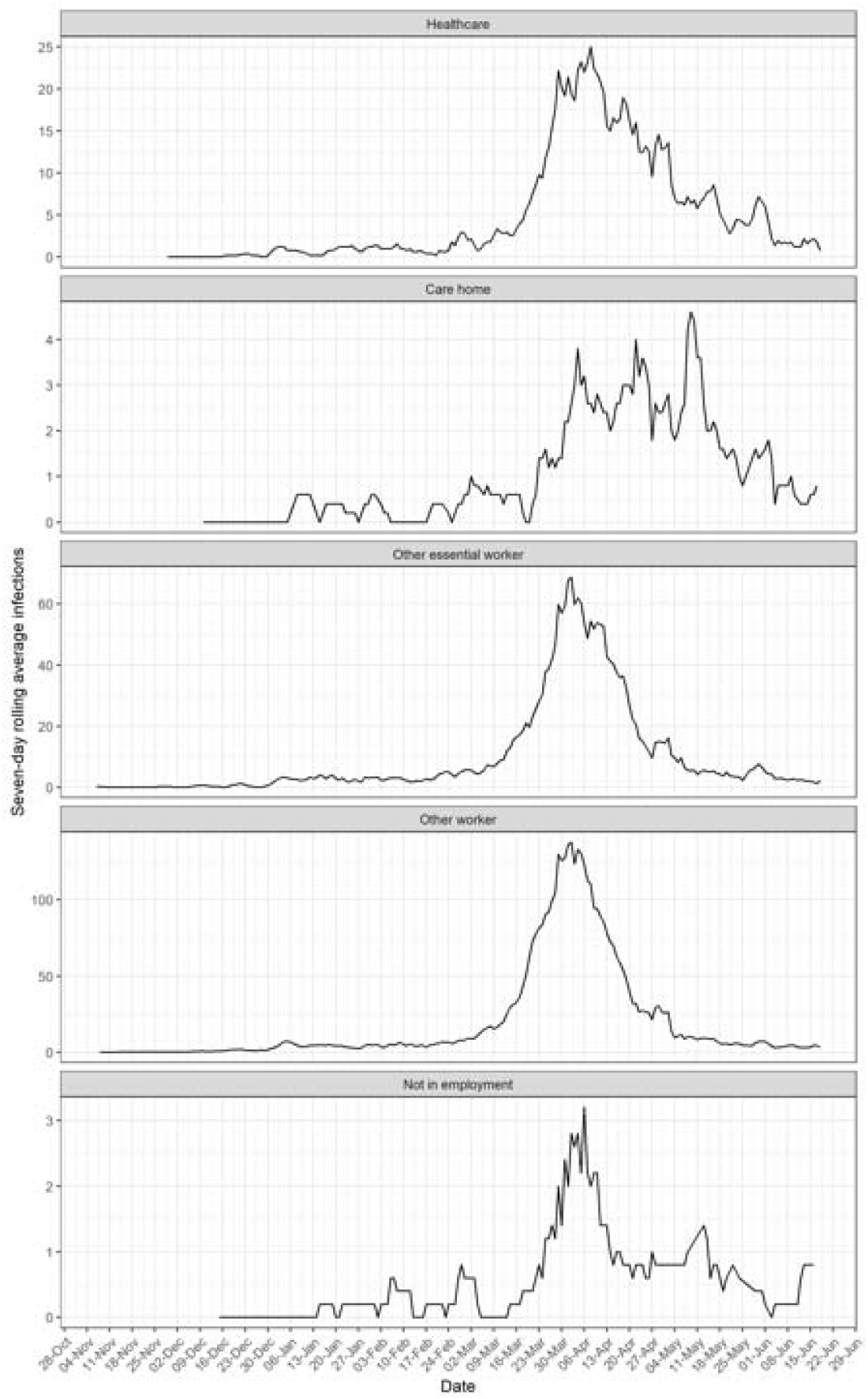
Epidemic curve reconstructed from reported date of onset from all three rounds by employment type, to June 2020. Seven-day rolling average of number of infections (by onset date) in participants testing positive for antibodies and who reported a date of onset for symptoms of COVID19 by employment type. Healthcare worker includes those with and without direct patient contact; care home worker includes those with and without direct client contact; other essential worker as defined by the UK Government https://www.gov.uk/guidance/coronavirus-covid-19-getting-tested#essential-workers includes those in emergency services, essential public services, transport and education; other worker includes workers not working in health or social care or on the UK Government list of essential workers.’

To check for consistency between rounds we compared the sensitivity cut-off points between the Fortress LFIA batches used in Round 1 and Rounds 2 and 3 using serial dilutions of sera from 10 PCR-confirmed SARS-CoV-2 infected individuals, and found a high level of consistency (Supplementary appendix figure 2). In laboratory-based assays using sera from health care workers who had recovered from SARS CoV2 infection, we found that a positive result on the LFIA used in the REACT 2 antibody prevalence study was associated with a higher titre of neutralising antibody. Sera that scored positive in the LFIA had a median neutralization titre of 40 which was significantly (P<0.0001) higher than those that scored negative with a median of zero (Figure 6).

**Figure 6:**
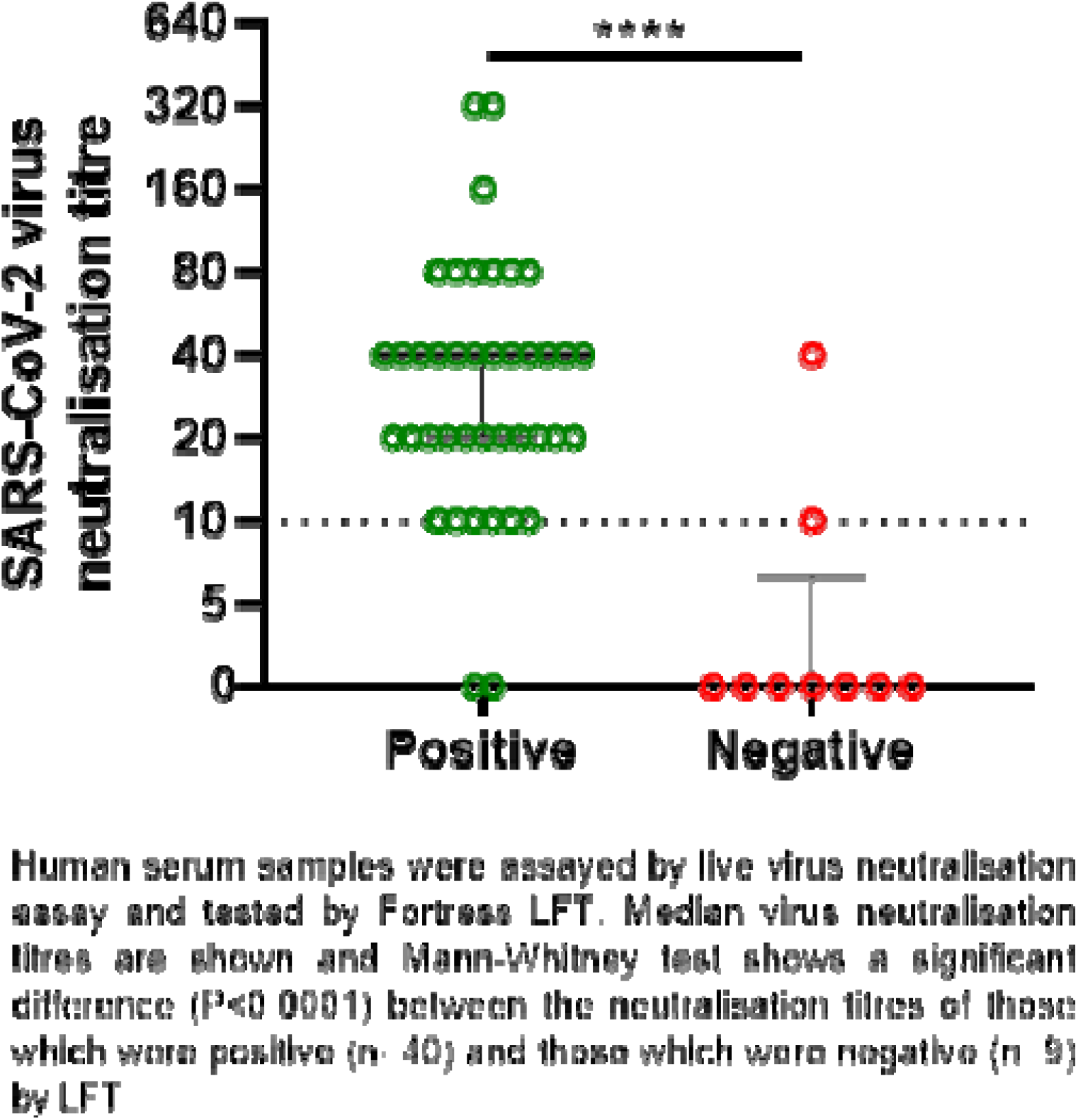
Association of LFIA result with virus microneutralisation titre in 49 healthcare workers with PCR-confirmed SARS-CoV-2 infection.

## Discussion

We observe a significant decline in the proportion of the population with detectable antibodies over three rounds of national surveillance, using a self-administered lateral flow test, 12, 18 and 24 weeks after the first peak of infections in England. This is consistent with evidence that immunity to seasonal coronaviruses declines over 6 to 12 months after infection and emerging data on SARS-CoV-2 that also detected a decrease over time in antibody levels in individuals followed in longitudinal studies.^4,5,9^ We observed clear differences in rates of decline between groups, for example those reporting SARS-CoV-2 infection based on PCR versus those without a history of COVID-19. In some groups with continued exposure risks no change in prevalence was seen (e.g. healthcare workers).

The relevance of antibody waning for the potential for reinfection by SARS CoV-2 is currently not resolved.^19,20^ During any antibody response to an acute pathogen, some level of antibody waning in the months following infection is expected as short lived plasma cells die. Low levels of affinity-matured antibody usually continues to be produced by long-lived plasma cells, and may be sufficient to maintain levels of antibody that confer immunity. Indeed for some pathogens such as measles, influenza and rhinovirus, antibodies can be detected for many years after infection. However the situation for coronaviruses is less clear. Human challenge studies showed a more profound waning of serum and nasal antibody over one year following coronavirus challenge than was seen for volunteers challenged with rhinovirus. At one year, re-infection with the seasonal coronavirus was observed whereas volunteers who retained antibodies following rhinovirus infection displayed sterilizing immunity. ^21,22^

Moreover modelling shows that waning immunity can explain the 1-2 year periodicity of reinfections with seasonal coronaviruses.^23^ Although reports of reinfection with SARS-CoV2 have been limited to date,^24^ this is in part because definitive evidence of reinfection requires sequencing of virus at two time points, which is rarely available in practice. In addition, asymptomatic testing is not yet widespread in many countries and thus mild or asymptomatic reinfections will go undetected. Understanding the ongoing risks of reinfection for the population is key to understanding the future course of the epidemic.

It is widely thought that titres of anti-Spike (S) antibodies which target the receptor binding domain (RBD, associated with cell entry) correlate with protection from reinfection. ^25,26^The lateral flow test used for this study detects antibodies against the spike protein (anti-S), but is qualitative rather than quantitative, and the threshold of detection is not stated in manufacturer’s instructions. We tested serial dilutions of known positive sera in the LFIA and confirmed that for each of the sera there was a different dilution after which the LFIA no longer yielded a positive band (Supplementary Appendix Figure 2). This demonstrates that, as antibody wanes from a population with a diverse mixture of starting titres, gradually the proportion of positive individual tests will decline. Our data in Figure 6 suggest the threshold for detection of antibody in sera with the LFIA corresponds to serum endpoint titres that score between 1:10 and 1:40 in a live virus microneutralisation assay. We cannot know at this time how this relates to the level of antibody that confers protection from infection, though studies in non-human primates vaccinated with an array of vaccines that conferred varying levels of immunity, suggest these may be similar levels to those required for protection.^27^ The relevant thresholds for protection in humans who are naturally exposed to virus remain to be defined and will continue to be informed by detailed studies of outbreaks.^28^ In addition it is currently not clear what contribution T cell immunity and memory responses will play in protective immunity during re-exposure. As such, it is not possible to say with certainty that the loss of antibody positivity in the LFIA would correlate with an increased risk of an individual being reinfected. However, at a population level, the waning we have observed may indicate an overall decline in the level of population immunity.

The declining prevalence of antibodies raises the question as to the extent to which antibody prevalence estimated during round one of our study, approximately 3 months after the peak of the first wave, may have underestimated the total of those infected in the first wave in the UK. We reported a prevalence of 6.0% (95% CI: 5.8-6.1) from round one (20 June to 13 July 2020), implying that at least 3.36 (3.22, 3.51) million adults in England had been infected with SARS-CoV-2 and tested positive for antibodies.^13^ Descriptions of the decline following infection are variable, with a general consensus that IgG levels can remain high for 2-3 months before declining,^9,29^ but those with smaller initial antibody responses are likely to decline earlier.^9^ Decline may initially be rapid, before plateauing, but data on this are only now beginning to emerge. Our previous estimate of antibody prevalence was consistent with that from the smaller ONS survey which reports antibody prevalence declining from 7.4% (95% CI 5.6, 9.6) in May to 5.6% (5.0, 6.2) in September.^30^

Our study has limitations. It included non-overlapping random samples of the population, but it is possible that people who had been exposed to the virus were less likely to take part over time, which may have contributed to apparent population antibody waning. However, we had similar response rates across the three surveys, and for each round, we re-weighted the sample to be representative of the country as a whole. We adjusted for test characteristics (sensitivity, specificity) based on our evaluation in clinic-based tests among healthcare workers with confirmed infection, carried out before the first round,^16^ but changes in prevalence are unlikely to be a consequence of batch variation in tests. We compared the laboratory performance of the LFIAs used in rounds 1 and 2 (where we had seen the strongest decline in positive tests) and found no difference between the two rounds. We also did not detect differences in ability of participants to use the LFIA (indeed, failure rates were lower in later rounds compared to earlier ones). The characteristics of the test mean that results are not appropriate for clinical use in individuals and participants are advised not to change their behaviour based on the result. However, as participants are not blind to the results of their LFIA it is possible that this may have introduced bias into their questionnaire response, but this should not have affected our observation of declining prevalence over time.

In summary, our findings provide evidence of variable waning in antibody positivity over time based on detectable IgG antibodies using a lateral flow assay. These data suggest the possibility of decreasing population immunity and increasing risk of reinfection as detectable antibodies decline in the population.

## Data Availability

The original datasets generated or analysed, or both, during this study are not publicly available because of governance restrictions and the identifiable nature of the data.

## Acknowledgements

We thank key collaborators on this work -- Ipsos MORI: Stephen Finlay, John Kennedy, Kevin Pickering, Duncan Peskett, Sam Clemens and Kelly Beaver; Institute of Global Health Innovation at Imperial College: Gianluca Fontana, Dr Hutan Ashrafian, Sutha Satkunarajah and Lenny Naar; Imperial College Healthcare NHS Trust: Robert Klaber; the Patient Experience Research Centre and the REACT Public Advisory Panel; NHS Digital for access to the NHS Register.

HW is a NIHR Senior Investigator and acknowledges support from NIHR Biomedical Research Centre of Imperial College NHS Trust, NIHR School of Public Health Research, NIHR Applied Research Collaborative North West London, Wellcome Trust 205456/Z/16/Z.

GC is supported by an NIHR Professorship. WSB is the Action Medical Research Professor, AD is an NIHR senior investigator and DA is an Emeritus NIHR Senior Investigator.

SR acknowledges support from MRC Centre for Global Infectious Disease Analysis, National Institute for Health Research (NIHR) Health Protection Research Unit (HPRU), Wellcome Trust (200861/Z/16/Z, 200187/Z/15/Z), and Centres for Disease Control and Prevention (US, U01CK0005-01-02)

PE is Director of the MRC Centre for Environment and Health (MR/L01341X/1, MR/S019669/1). PE acknowledges support from the NIHR Imperial Biomedical Research Centre and the NIHR HPRUs in Environmental Exposures and Health and Chemical and Radiation Threats and Hazards, the British Heart Foundation Centre for Research Excellence at Imperial College London (RE/18/4/34215) and the UK Dementia Research Institute at Imperial (MC_PC_17114).

We thank the Huo Family Foundation for support of our work on COVID-19.

