## Supplementary methods and figures for "Declining prevalence of antibody positivity to SARS-CoV-2: a community study of 365,000 adults"

### 1. **Supplementary Methods**

#### 1.1 Table S1: Response rates

|  | Round 1 20 Jun – 13 July | % of all invited | Round 2 31 Jul – 13 Aug | % of all invited | Round 3 15-28 Sept | % of all invited | **All rounds** | **% of all invited** |
| --- | --- | --- | --- | --- | --- | --- | --- | --- |
| Invitation letters sent | 314996 |  | 344737 |  | 560385 |  | **1220118** |  |
| Registered* | 126143 | 40.0% | 131596 | 38.2% | 201720 | 36.0% | **459459** | **37.7%** |
| Valid result | 99908 | 31.7% | 105829 | 30.7% | 159367 | 28.4% | **365104** | **29.9%** |

** Registration was stopped when target reached approx. 120,000 in rounds 1 and 2, 200,000 in round 3 to ensure sufficient kits would be available, and therefore overall response rate is artificially capped.*

#### 1.2 Weighting Strategy

The weighting approach used rim weighting to adjust to population estimates of: age by sex; Index of Multiple Deprivation (IMD) deciles; Local Authority (LA) counts; ethnic group.

The age by gender and LA counts were extracted from the ONS mid-year population estimates (1), the ethnic group counts from the Labour Force Survey (Annual Population Survey) (2), and the IMD deciles profile were derived from the original anonymous population sampling frame from NHS digital. To allow for the different sources of population estimates, the rim weighting was carried out on the proportions rather than population totals.

Age was grouped into seven categories: 18 to 24; 25 to 34; 35 to 44; 45 to 54; 55 to 64; 65 to 74; 75 or older, giving 14 age-sex categories.

The reported ethnicity was grouped into nine categories: white; mixed / multiple ethnic groups; Indian; Pakistani; Bangladeshi; Chinese; any other Asian background; black African / Caribbean / other; and any other ethnic group or missing.

The rim weighting was carried out in two stages. At the first stage, the sample was weighted to LA counts and age by gender groups only. This put the sample back into the correct proportion for LAs which corrects for the disproportionate sampling and differential non-response. In the same stage, the age and gender groups were also adjusted to make sure that the final weighted profile was as close to the population as possible.

The second stage of rim weighting adjusted to all four measures, using the first stage weights as the starting weights. The adjustment factor between the first and second stage weights were trimmed at the 1^st^ and 99^th^ percentiles to dampen the extreme weights which improves efficiency. The final weights were calculated as the first stage weights multiplied by the trimmed adjustment factor for the second stage.

### 2. **Supplementary Figures**

#### 2.1 Figure S1: Map of prevalence of Sars-CoV-2 antibody by lower tier local authority, England over three rounds

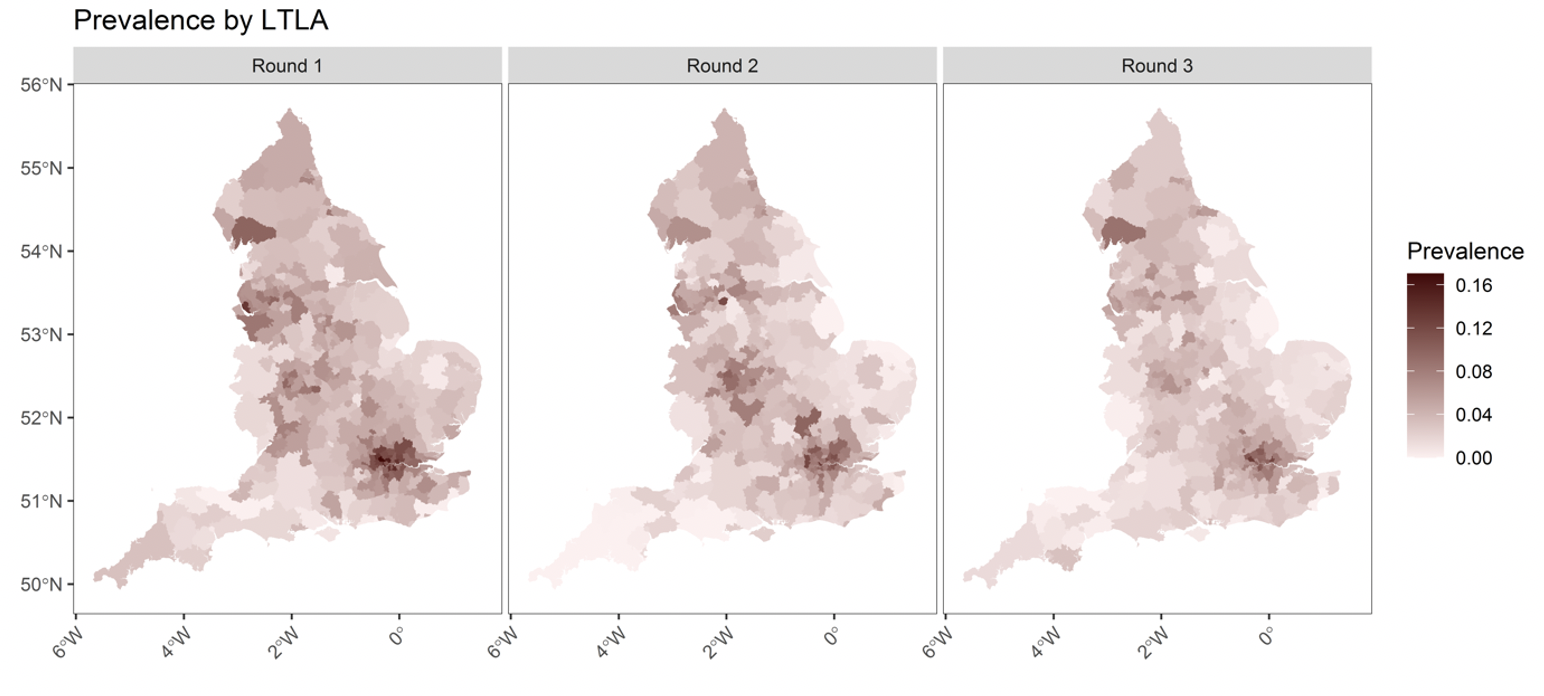

Legend: the prevalence of antibodies in each LTLA area by round of study.

#### 2.2 Figure S2: Comparison of the sensitivity cut-off points between the Fortress LFIAs used in Round 1 (R1) and Rounds 2 and 3 (R2)

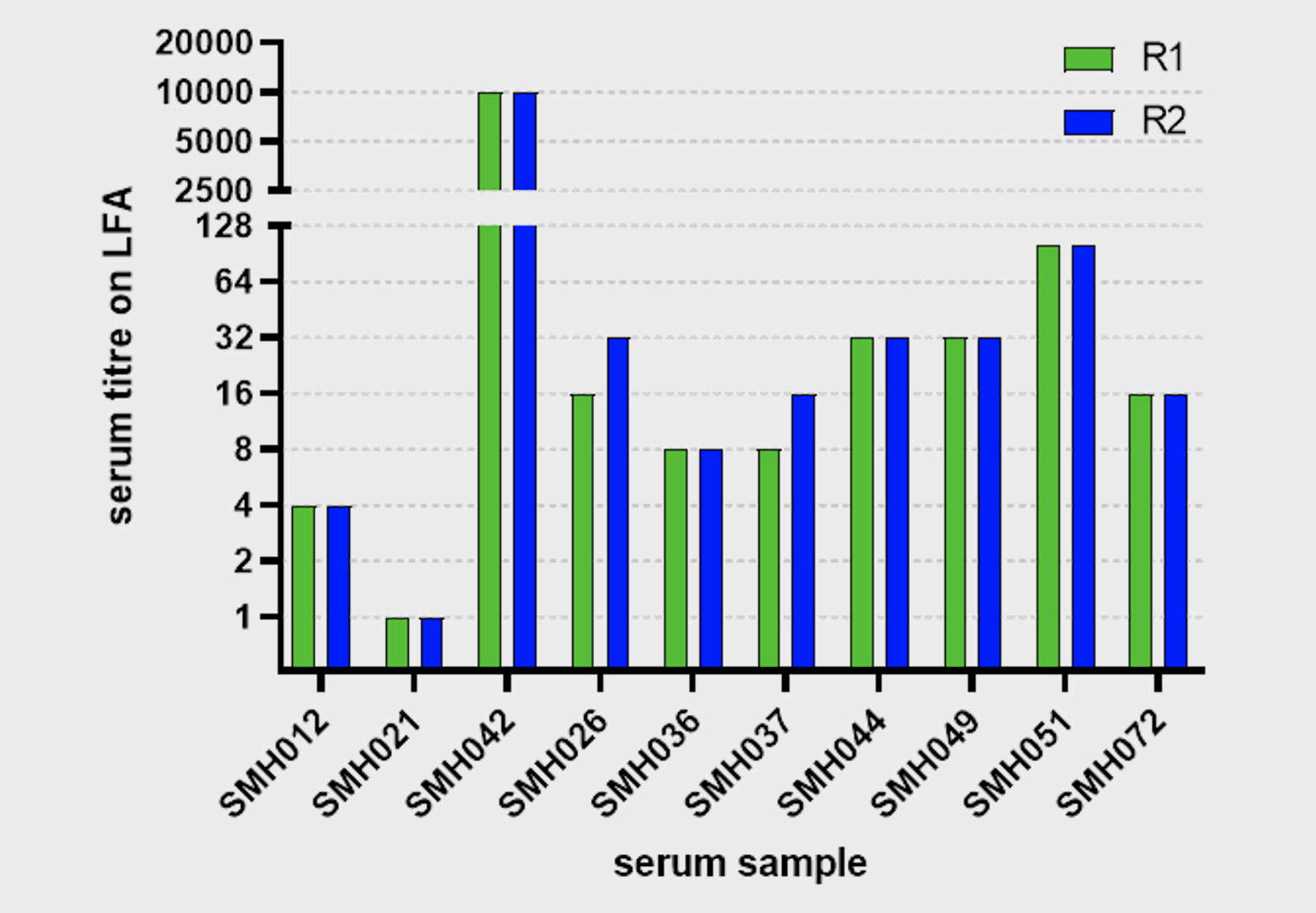

**Legend:**  Comparison of the sensitivity cut-off points between the Fortress LFIAs used in Round 1 (R1) and Rounds 2 and 3 (R2) of REACT2 Study 5 using 2 and 10-fold dilutions of sera from 10 PCR-confirmed SARS-CoV-2 infected individuals

### 3. **References**

1.	Office_for_National_Statistics. Population estimates for the UK, England and Wales, Scotland and Northern Ireland: mid-2019 2020 [Available from: [www.ons.gov.uk/releases/populationestimatesfortheukenglandandwalesscotlandandnorthernirelandmid2019](https://imperiallondon-my.sharepoint.com/personal/ca1411_ic_ac_uk/Documents/Nature/www.ons.gov.uk/releases/populationestimatesfortheukenglandandwalesscotlandandnorthernirelandmid2019).

2.	Office_for_National_Statistics. Annual Population Survey/Labour Force Survey 2020 [Available from: <https://www.nomisweb.co.uk/sources/aps>.
